# Trends in non-daily cigarette smoking in England, 2006–2024

**DOI:** 10.1101/2024.07.17.24310556

**Authors:** Sarah E. Jackson, Jamie Brown, Lion Shahab, Sharon Cox

## Abstract

**Background:** Cigarette smoking is incredibly harmful, even for people who do not smoke every day. This study aimed to estimate trends in non-daily smoking in England between 2006 and 2024, how these differed across population subgroups, and to explore changes in the profile of non-daily smokers in terms of their sociodemographic and smoking characteristics and vaping and alcohol consumption.

**Methods:** Data were collected monthly between November-2006 and April-2024 as part of a nationally-representative, repeat cross-sectional survey of adults (≥18y; *n*=353,711). We used logistic regression to estimate associations between survey wave and non-daily smoking and used descriptive statistics to characterise the profile of non-daily smokers across 3-year periods.

**Results:** The proportion of cigarette smokers who smoked non-daily was relatively stable between November-2006 and November-2013, at an average of 10.5% [10.1-10.9%], then increased to 27.2% [26.0-28.4%] (4.0% [3.7-4.2%] of adults) by April-2024. This increase was particularly pronounced among younger adults (e.g., reaching 52.8%, 20.4%, and 14.4% of 18-, 45-, and 65-year-old cigarette smokers by April-2024) and those who vape (reaching 34.2% vs. 23.1% among non-vapers). Over time, there were reductions in non-daily smokers’ mean weekly cigarette consumption (from 34.3 in 2006-09 to 21.1 in 2021-24), urges to smoke (e.g., the proportion reporting no urges increased from 29.2% to 38.0%), and motivation to stop smoking (e.g., the proportion highly motivated to quit within the next three months decreased from 30.8% to 21.0%).

**Conclusions:** An increasing proportion of adults in England who smoke cigarettes do not smoke every day, particularly younger adults. Although non-daily smokers report smoking fewer cigarettes and weaker urges to smoke than they used to, which may make it easier for them to stop smoking, they appear to be decreasingly motivated to quit.

## Introduction

Cigarette smoking is uniquely harmful. It substantially increases the risks of disease, disability, and premature death^1–4^ – even for people who do not smoke every day.^5–9^ In England, most adults who smoke report doing so multiple times a day, with the average smoker consuming around 11 cigarettes each day.^10^ However, non-daily smoking (sometimes referred to as intermittent or occasional smoking, or ‘chipping’^11^) appears to have become more common in recent years.^10,12^ For example, representative population surveys conducted in 2008 and 2017 showed the proportion of current smokers in England who do not smoke every day increased from 9.1% to 13.4% over this period.^12^ A similar pattern has been documented in other countries, such as the US, albeit over a much longer time period.^13^ A recent study reported a further increase up to 2023 in England^10^ but it is not clear the extent to which these increases have differed by key sociodemographic groups and smoking and vaping characteristics.

Non-daily smokers often minimise the health effects of their smoking^14,15^ and many do not even consider themselves smokers.^16,17^ There may be a misperception that non-daily smoking is less harmful or addictive than daily smoking, but even non-daily smoking carries substantial risks relative to not smoking,^5–7,9^ including similar cardiovascular risks as daily smoking.^9^ In terms of dependence and quitting, studies have found non-daily smokers tend to report lower levels of addiction than daily smokers,^15,18^ being more ready to quit,^16^ and having higher confidence in their ability to quit.^17^ However, tobacco smoking is so addictive, and often bound to social and environment contexts, that even low-level smokers experience a loss of autonomy over their smoking behaviour,^19,20^ meaning they also struggle to quit.^21,22^ As such, although they are more likely to succeed in stopping smoking than daily smokers, the majority of quit attempts made by non-daily smokers are unsuccessful in the long-term.^21,22^ They also tend to be less likely than daily smokers to receive advice or support for smoking cessation from healthcare professionals.^23,24^

It is important to understand more about non-daily smoking in England to inform interventions to further reduce smoking in the population, including public health messaging and provision of cessation support. While studies show an overall increase in the proportion of smokers reporting non-daily use, it is not clear how far these changes have occurred across different subgroups of smokers. For example, there may be differences by sociodemographic characteristics. Cross-sectional studies have generally found that non-daily smokers tend to be younger than daily smokers and are more likely to be female and from more advantaged socioeconomic groups.^16,22,25^ Associations with alcohol consumption have also been documented, with non-daily smokers more likely than daily smokers to drink excessively and to smoke cigarettes while under the influence of alcohol.^26–29^ In addition, many smokers who vape (‘dual users’) report using e-cigarettes to quit or cut down on smoking.^30^ E-cigarettes offer a harm reduction alternative to combustible tobacco^31^ and are an effective quit aid,^32^ but users must completely switch to e-cigarettes for the greatest health benefits. Recent data suggest there has been a greater decline in the average number of cigarettes smoked per day among smokers who vape compared with those who do not,^10^ which may have led to larger increases in non-daily smoking among dual users. Up-to-date information on the profile of non-daily smokers is needed, to understand what this group currently looks like in terms of their sociodemographic characteristics; their drinking, vaping, and smoking behaviour; and their intentions to stop smoking.

The Smoking Toolkit Study, a repeat cross-sectional household survey in England, has been collecting data on non-daily smoking from representative samples of adults each month since November 2006. This study used these data to provide a detailed update on the extent to which changes in trends in non-daily smoking in England between 2006 and 2024 have differed by age, gender, occupational social grade, vaping status, and level of alcohol consumption. It also explored changes in the profile of non-daily smokers over this period.

## Methods

### Pre-registration

The study protocol, research questions, and analysis plan were pre-registered on Open Science Framework (https://osf.io/xw8du/).

### Design

The Smoking Toolkit Study uses a hybrid of random probability and simple quota sampling to select a new sample of approximately 1,700 adults (≥16y) each month. ^33,34^ Comparisons with other national surveys and sales data indicate the survey achieves nationally representative estimates of key variables including sociodemographic characteristics, smoking prevalence, and cigarette consumption.^33,35^

Data were collected via face-to-face interviews up to the start of the pandemic. No data were collected in March 2020 and interviews were conducted via telephone from April 2020 onwards. The two modes of data collection show good comparability on key sociodemographic and smoking variables.^36,37^ The sample excluded 16- and 17-year-olds between April 2020 and December 2021.

We analysed data collected between November 2006 (the first wave of data collected) and April 2024 (the most recent data available at the time of analysis). We restricted the sample to participants aged ≥18 years for consistency across the time series.

### Measures

Full details of the measures are provided in the study protocol (https://osf.io/xw8du/).

Smoking status was assessed by asking participants which of the following best applied to them: (a) I smoke cigarettes (including hand-rolled) every day; (b) I smoke cigarettes (including hand-rolled), but not every day; (c) I do not smoke cigarettes at all, but I do smoke tobacco of some kind (e.g., pipe, cigar or shisha); (d) I have stopped smoking completely in the last year; (e) I stopped smoking completely more than a year ago; (f) I have never been a smoker (i.e., smoked for a year or more). Those who responded (a) or (b) were considered current cigarette smokers. Those who responded (b) were considered non-daily smokers.

Participants reported their age, gender, and occupational social grade (ABC1 includes managerial, professional, and upper supervisory occupations / C2DE includes manual routine, semi-routine, lower supervisory, state pension, and long-term unemployed). Past-6-month alcohol consumption was assessed with the three-item AUDIT-C (range: 0–12, higher scores indicate higher levels of consumption).^38^ Vaping status was categorised as current vaper vs. non-vaper.

Among current cigarette smokers, we also recorded the number of cigarettes usually smoked per week, the main type of cigarettes smoked (manufactured/hand-rolled), strength of urges to smoke over the past 24 hours,^39^ motivation to stop smoking,^40^ and past-year quit attempts.

Some variables were not assessed across the entire period: the main type of cigarettes smoked was assessed from January 2008, alcohol consumption from March 2014, and vaping status from April 2011 among cigarette smokers and from October 2013 among all adults. Analyses using these variables were therefore limited to the period when data were available.

### Statistical analysis

Data were analysed using R v.4.2.1. The Smoking Toolkit Study uses raking to weight the sample to match the population in England.^33^ The following analyses used weighted data. We excluded participants with missing data on non-daily smoking. Missing cases on other variables were excluded on a per-analysis basis.

#### Trends in non-daily vaping by sociodemographic, vaping, and drinking subgroups

Trends in the proportion of (i) adults and (ii) cigarette smokers reporting non-daily smoking over the study period were analysed using logistic regression, with non-daily smoking as the outcome and time (survey month) modelled using restricted cubic splines. This allowed for flexible and non-linear changes over time, while avoiding categorisation.

To explore moderation of trends among cigarette smokers by age, gender, occupational social grade, vaping status, and level of alcohol consumption, we repeated the model including the interaction between the moderator of interest and time – thus allowing for time trends to differ across subgroups. Each of the interactions was tested in a separate model. Age and alcohol consumption (AUDIT-C) were modelled using restricted cubic splines with three knots (placed at the 5, 50, and 95th percentiles), to allow for non-linear relationships with non-daily smoking.

For each analysis (i.e., overall trends and interactions, among adults and among cigarette smokers), we compared models with survey wave analysed using restricted cubic splines with three, four, and five knots (sufficient to accurately model trends across years without overfitting) using the Akaike Information Criterion (AIC). The best fitting model was selected as the model with the lowest AIC or the simplest model within two AIC units (see **Table S1** for details).

We used predicted estimates from the best fitting models to plot non-daily smoking prevalence over the study period. We reported prevalence ratios (PRs) for changes from November 2006 (or the first month of data available, for analyses by vaping status and alcohol consumption) to April 2024 alongside 95% confidence intervals (CIs) calculated using bootstrapping. We also plotted unmodelled datapoints showing the prevalence of daily and non-daily smoking by survey year, to provide context.

In an unplanned analysis, we repeated the interaction with vaping status stratified by age group, and the interaction with age stratified by vaping status, to explore these associations among cigarette smokers in more detail.

#### Changes in the profile of non-daily smokers

We used descriptive statistics to compare the sociodemographic, vaping, drinking, and smoking profiles of non-daily cigarette smokers over the study period. Time was categorised in three-year periods (November 2006 to October 2009, etc.) to provide adequate sample sizes for this analysis.

In an unplanned analysis, we explored changes in the strength of urges to smoke among non-daily cigarette smokers, stratified by vaping status.

## Results

Data were collected from 354,480 participants aged ≥18y in England between November 2006 and April 2024. We excluded 769 (0.2%) who did not respond to the question on smoking status, leaving a sample of 353,711 adults for analysis (unweighted mean [SD] age 49.1 [19.0] years; 181,318 [51.3%] women), of whom 66,792 (18.9%) reported current cigarette smoking (unweighted mean [SD] age 42.9 [16.8] years; 32,795 [49.1%] women).

### Trends in non-daily smoking

**Figure 1** shows unmodelled datapoints and modelled trends in non-daily cigarette smoking among all adults and among cigarette smokers. **Figure 2** shows modelled trends in non-daily smoking among subgroups of adults and cigarette smokers. **Table 1** summarises modelled estimates of changes in non-daily smoking from the start to the end of the study period.

**Figure 1.**
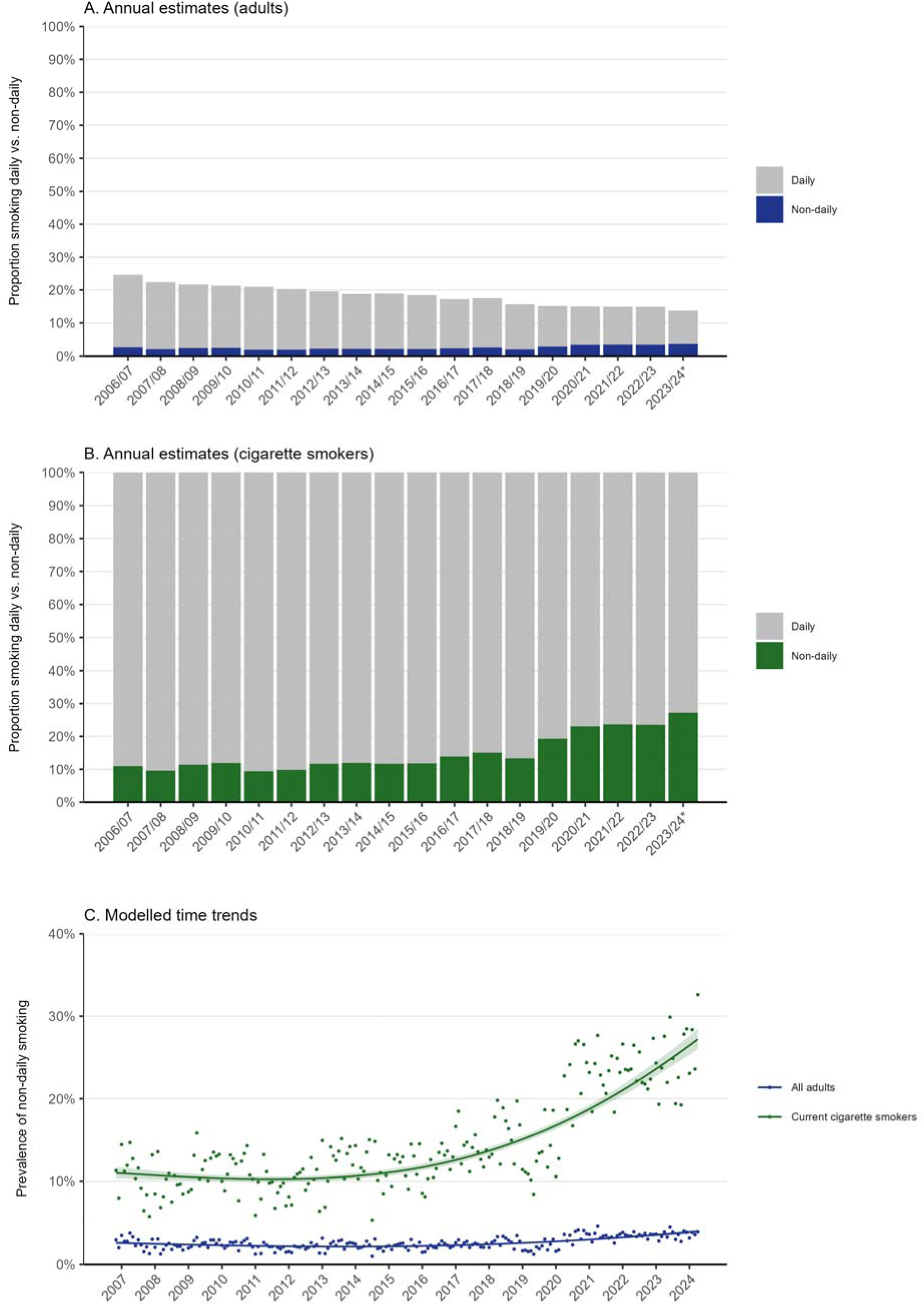
Prevalence of non-daily smoking among adults (≥18y) in England, 2006 to 2024. Panels A and B show weighted data aggregated by year, among adults and cigarette smokers respectively. Bars represent the proportions smoking daily and non-daily. Data are aggregated across 12-month periods (November to October). *Data for 2023/24 are based on November to April only. Panel C shows modelled time trends among adults and cigarette smokers. Lines represent modelled weighted prevalence of non-daily smoking by monthly survey wave, modelled non-linearly using restricted cubic splines (best fitting models; see **Table S1** for model selection). Shaded bands represent 95% confidence intervals. Points represent the unmodelled weighted proportion by month. Unweighted sample sizes: adults *n*=353,711, cigarette smokers *n*=66,792.

**Figure 2.**
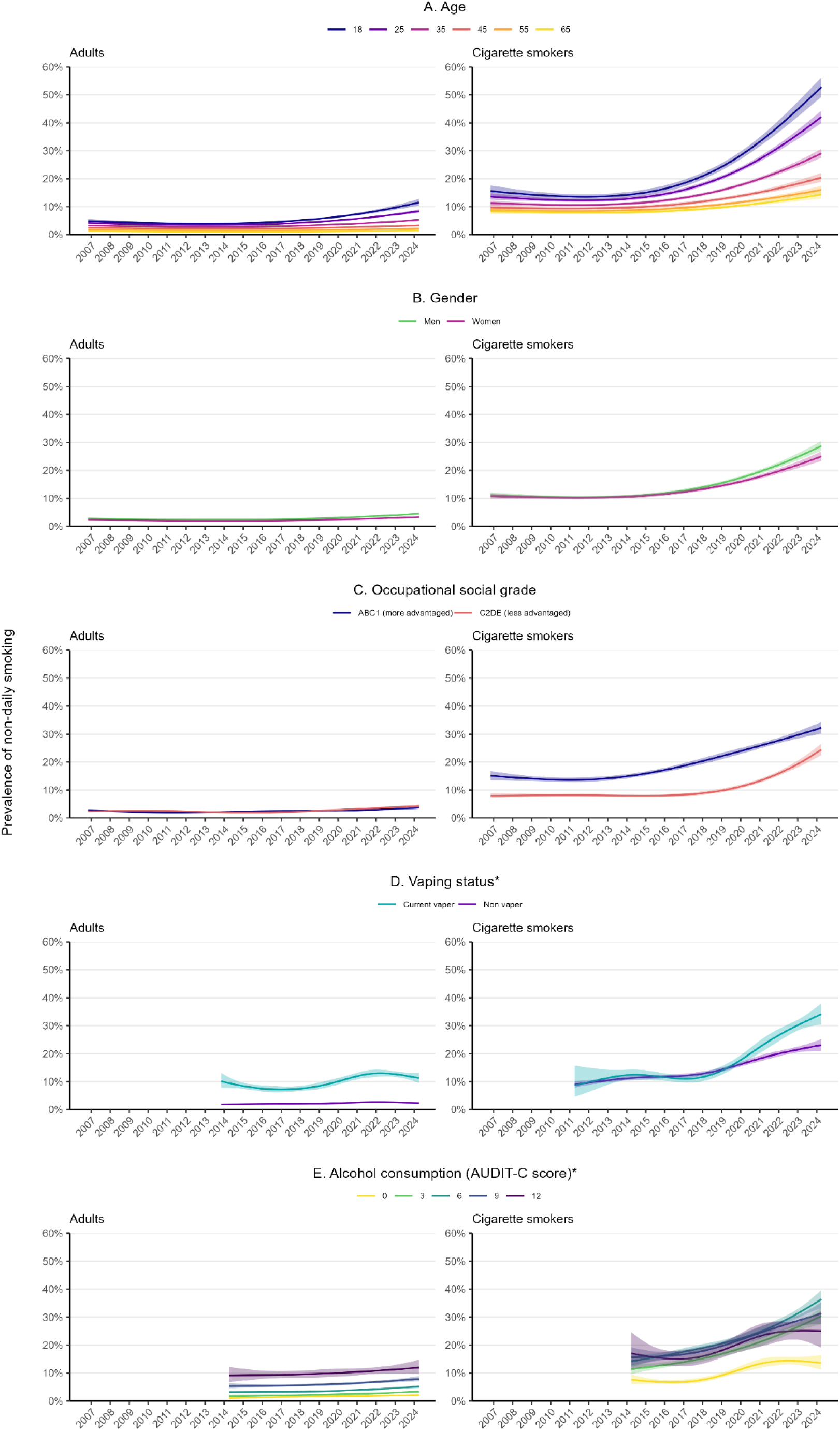
Trends in non-daily smoking among subgroups of adults (≥18y) and adult cigarette smokers in England, 2006 to 2024. Lines represent the modelled weighted proportion by monthly survey wave (modelled non-linearly using restricted cubic splines; best fitting models, see **Table S1** for model selection) and (A) age, (B) gender, (C) occupational social grade, (D) vaping status, and (E) level of alcohol consumption. Shaded bands represent 95% confidence intervals. *Data on vaping status were only available for cigarette smokers from April 2011 and for all adults from October 2013 and data on alcohol consumption from March 2014. Unweighted sample sizes (adults): trends by age and occupational social grade *n*=353,711; trend by gender *n*=352,924; trend by vaping status *n*=206,394; trend by level of alcohol consumption *n*=195,235. Unweighted sample sizes (cigarette smokers): trends by age and occupational social grade *n*=66,792; trend by gender *n*=66,614; trend by vaping status *n*=42,259; trend by level of alcohol consumption *n*=30,785.

**Table 1.**
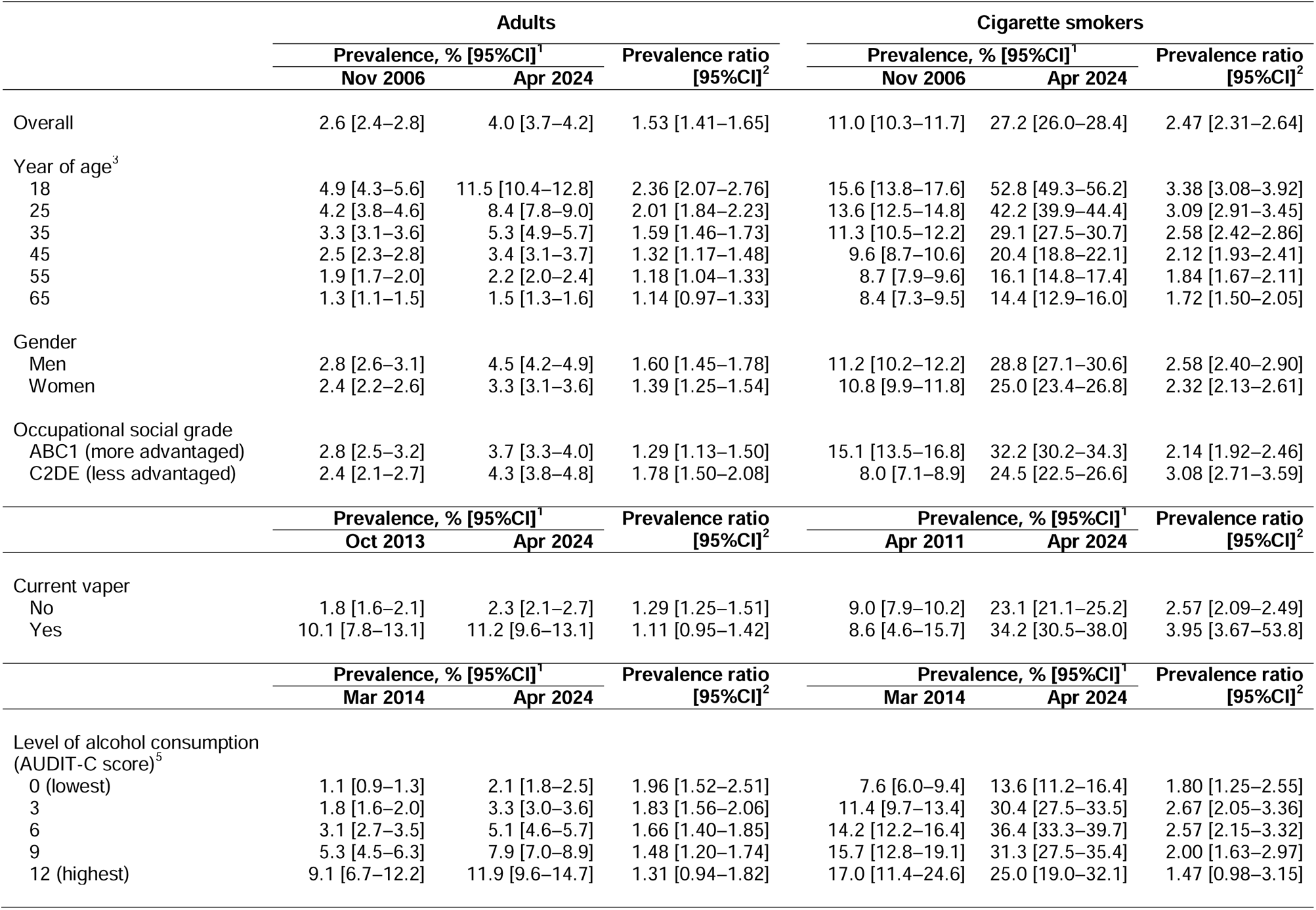
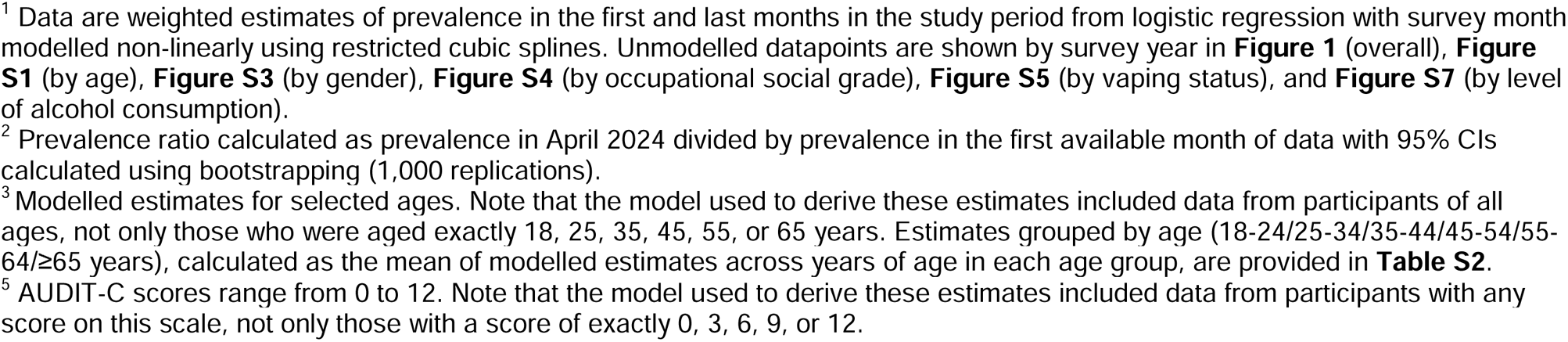
Modelled estimates of the prevalence of non-daily smoking among adults in the first available monthly wave and in April 2024.

|  | Adults |  |  | Cigarette smokers |  |  |
| --- | --- | --- | --- | --- | --- | --- |
|  | Prevalence, % [95%CI]<sup>1</sup> |  | Prevalence ratio<br>[95%CI]<sup>2</sup> | Prevalence, % [95%CI]<sup>1</sup> |  | Prevalence ratio<br>[95%CI]<sup>2</sup> |
|  | Nov 2006 | Apr 2024 |  | Nov 2006 | Apr 2024 |  |
| Overall | 2.6 [2.4–2.8] | 4.0 [3.7–4.2] | 1.53 [1.41–1.65] | 11.0 [10.3–11.7] | 27.2 [26.0–28.4] | 2.47 [2.31–2.64] |
| Year of age<sup>3</sup> |  |  |  |  |  |  |
| 18 | 4.9 [4.3–5.6] | 11.5 [10.4–12.8] | 2.36 [2.07–2.76] | 15.6 [13.8–17.6] | 52.8 [49.3–56.2] | 3.38 [3.08–3.92] |
| 25 | 4.2 [3.8–4.6] | 8.4 [7.8–9.0] | 2.01 [1.84–2.23] | 13.6 [12.5–14.8] | 42.2 [39.9–44.4] | 3.09 [2.91–3.45] |
| 35 | 3.3 [3.1–3.6] | 5.3 [4.9–5.7] | 1.59 [1.46–1.73] | 11.3 [10.5–12.2] | 29.1 [27.5–30.7] | 2.58 [2.42–2.86] |
| 45 | 2.5 [2.3–2.8] | 3.4 [3.1–3.7] | 1.32 [1.17–1.48] | 9.6 [8.7–10.6] | 20.4 [18.8–22.1] | 2.12 [1.93–2.41] |
| 55 | 1.9 [1.7–2.0] | 2.2 [2.0–2.4] | 1.18 [1.04–1.33] | 8.7 [7.9–9.6] | 16.1 [14.8–17.4] | 1.84 [1.67–2.11] |
| 65 | 1.3 [1.1–1.5] | 1.5 [1.3–1.6] | 1.14 [0.97–1.33] | 8.4 [7.3–9.5] | 14.4 [12.9–16.0] | 1.72 [1.50–2.05] |
| Gender |  |  |  |  |  |  |
| Men | 2.8 [2.6–3.1] | 4.5 [4.2–4.9] | 1.60 [1.45–1.78] | 11.2 [10.2–12.2] | 28.8 [27.1–30.6] | 2.58 [2.40–2.90] |
| Women | 2.4 [2.2–2.6] | 3.3 [3.1–3.6] | 1.39 [1.25–1.54] | 10.8 [9.9–11.8] | 25.0 [23.4–26.8] | 2.32 [2.13–2.61] |
| Occupational social grade |  |  |  |  |  |  |
| ABC1 (more advantaged) | 2.8 [2.5–3.2] | 3.7 [3.3–4.0] | 1.29 [1.13–1.50] | 15.1 [13.5–16.8] | 32.2 [30.2–34.3] | 2.14 [1.92–2.46] |
| C2DE (less advantaged) | 2.4 [2.1–2.7] | 4.3 [3.8–4.8] | 1.78 [1.50–2.08] | 8.0 [7.1–8.9] | 24.5 [22.5–26.6] | 3.08 [2.71–3.59] |
|  | Prevalence, % [95%CI]<sup>1</sup> |  | Prevalence ratio<br>[95%CI]<sup>2</sup> | Prevalence, % [95%CI]<sup>1</sup> |  | Prevalence ratio<br>[95%CI]<sup>2</sup> |
|  | Oct 2013 | Apr 2024 |  | Apr 2011 | Apr 2024 |  |
| Current vaper |  |  |  |  |  |  |
| No | 1.8 [1.6–2.1] | 2.3 [2.1–2.7] | 1.29 [1.25–1.51] | 9.0 [7.9–10.2] | 23.1 [21.1–25.2] | 2.57 [2.09–2.49] |
| Yes | 10.1 [7.8–13.1] | 11.2 [9.6–13.1] | 1.11 [0.95–1.42] | 8.6 [4.6–15.7] | 34.2 [30.5–38.0] | 3.95 [3.67–53.8] |
|  | Prevalence, % [95%CI]<sup>1</sup> |  | Prevalence ratio<br>[95%CI]<sup>2</sup> | Prevalence, % [95%CI]<sup>1</sup> |  | Prevalence ratio<br>[95%CI]<sup>2</sup> |
|  | Mar 2014 | Apr 2024 |  | Mar 2014 | Apr 2024 |  |
| Level of alcohol consumption<br>(AUDIT-C score)<sup>5</sup> |  |  |  |  |  |  |
| 0 (lowest) | 1.1 [0.9–1.3] | 2.1 [1.8–2.5] | 1.96 [1.52–2.51] | 7.6 [6.0–9.4] | 13.6 [11.2–16.4] | 1.80 [1.25–2.55] |
| 3 | 1.8 [1.6–2.0] | 3.3 [3.0–3.6] | 1.83 [1.56–2.06] | 11.4 [9.7–13.4] | 30.4 [27.5–33.5] | 2.67 [2.05–3.36] |
| 6 | 3.1 [2.7–3.5] | 5.1 [4.6–5.7] | 1.66 [1.40–1.85] | 14.2 [12.2–16.4] | 36.4 [33.3–39.7] | 2.57 [2.15–3.32] |
| 9 | 5.3 [4.5–6.3] | 7.9 [7.0–8.9] | 1.48 [1.20–1.74] | 15.7 [12.8–19.1] | 31.3 [27.5–35.4] | 2.00 [1.63–2.97] |
| 12 (highest) | 9.1 [6.7–12.2] | 11.9 [9.6–14.7] | 1.31 [0.94–1.82] | 17.0 [11.4–24.6] | 25.0 [19.0–32.1] | 1.47 [0.98–3.15] |

<sup>1</sup> Data are weighted estimates of prevalence in the first and last months in the study period from logistic regression with survey month modelled non-linearly using restricted cubic splines. Unmodelled datapoints are shown by survey year in **Figure 1** (overall), **Figure S1** (by age), **Figure S3** (by gender), **Figure S4** (by occupational social grade), **Figure S5** (by vaping status), and **Figure S7** (by level of alcohol consumption).
<sup>2</sup> Prevalence ratio calculated as prevalence in April 2024 divided by prevalence in the first available month of data with 95% CIs calculated using bootstrapping (1,000 replications).
<sup>3</sup> Modelled estimates for selected ages. Note that the model used to derive these estimates included data from participants of all ages, not only those who were aged exactly 18, 25, 35, 45, 55, or 65 years. Estimates grouped by age (18-24/25-34/35-44/45-54/55-64/ $\geq 65$ years), calculated as the mean of modelled estimates across years of age in each age group, are provided in **Table S2**.
<sup>5</sup> AUDIT-C scores range from 0 to 12. Note that the model used to derive these estimates included data from participants with any score on this scale, not only those with a score of exactly 0, 3, 6, 9, or 12.

#### Trends among adults and adult cigarette smokers

Between November 2006 and April 2024, the prevalence of non-daily smoking among adults in England increased from 2.6% to 4.0% (PR=1.53 [1.41–1.65]; **Table 1**). This represented a shift towards more non-daily smoking among current cigarette smokers, from 11.0% to 27.2% (PR=2.47 [2.31–2.64]; **Table 1**). However, the overall prevalence of cigarette smoking (either daily or non-daily) decreased non-linearly across the period (from 24.6% [24.0–25.3%] in 2006/07 to 13.7% [12.9–14.5%] in 2023/24; **Figure 1A**).

The increase in non-daily smoking prevalence was non-linear (**Figure 1C**). Up to the end of 2013, the proportion of cigarette smokers who smoked non-daily was relatively stable between 10.3% and 11.0% (at an average of 10.5% [10.1–10.9%] between November 2006 and November 2013). Given that overall smoking prevalence decreased, this meant the prevalence of non-daily smoking decreased from 2.6% [2.4–2.8%] in November 2006 to a low of 2.2% [2.1– 2.2%] in November 2013. The proportion of cigarette smokers who smoked non-daily then increased across the remainder of the period, reaching 27.2% [26.0–28.4%] by April 2024. The pattern was similar among all adults, with prevalence increasing to 4.0% [3.7–4.2%] by April 2024.

#### Trends within subgroups of adults and adult cigarette smokers

The prevalence of non-daily smoking was consistently higher across the study period at younger ages. The increase in prevalence between November 2006 and April 2024 was also larger at younger than older ages (e.g., PR=3.38 [3.08–3.92] for 18-year-old cigarette smokers vs. PR=2.12 [1.93–2.41] for 45-year-olds and PR=1.72 [1.50–2.05] for 65-year-olds), meaning the inverse age gradient in non-daily smoking became more pronounced over time (e.g., reaching 52.8%, 20.4%, and 14.4% of 18-, 45-, and 65-year-old cigarette smokers by April 2024; **Table 1**, **Figure 2A**, **Figure S1**). This pattern of results was observed among cigarette smokers who did and did not vape (**Figure S2**).

The prevalence of non-daily smoking was similar in men and women across most of the period. The proportion of cigarette smokers who smoked non-daily was slightly higher among men than women by April 2024 (28.8% vs. 25.0%, respectively), but there was overlap in the 95%CIs around PRs for changes from the start to the end of the period suggesting the difference was uncertain (PR=2.58 [2.40–2.90] for men; PR=2.32 [2.13–2.61] for women; **Table 1**, **Figure 2B**, **Figure S3**).

The prevalence of non-daily smoking was similar by occupational social grade among all adults, but consistently higher among cigarette smokers from more advantaged compared with less advantaged social grades. The rise in non-daily smoking started several years earlier among cigarette smokers from more advantaged social grades but the relative change between November 2006 and April 2024 was more pronounced among those from less advantaged social grades (PR=3.08 [2.71–3.59] for C2DE vs. PR=2.14 [1.92–2.46] for ABC1; **Table 1**, **Figure 2C**, **Figure S4**).

The prevalence of non-daily smoking was consistently higher among adults who vaped than those who did not but was similar by vaping status among cigarette smokers up to 2019. The increase in prevalence of non-daily smoking between April 2011 and April 2024 was greater among cigarette smokers who vaped than those who did not (PR=3.95 [3.67–53.8] vs. PR=2.57 [2.09–2.49]) (**Table 1**, **Figure 2D**, **Figure S5**). This may be partly explained by younger cigarette smokers being more likely both to vape (e.g., 21.7% [20.6–22.7%] across the period among 18-24-year-olds vs. 12.5% [11.6–13.5%] among ≥65-year-olds) and to smoke non-daily: trends within age groups appeared similar by vaping status (**Figure S6**). However, while the inverse age gradient that increased over the period was also apparent in non-vapers, it was more pronounced among vapers (**Figure S2**).

The prevalence of non-daily smoking was consistently higher among adults who drank more heavily. Among cigarette smokers, the proportion who smoked non-daily was lower among those who did not drink at all (AUDIT-C = 0) but was similar among drinkers. There was no notable difference in the increase in non-daily smoking by alcohol consumption (**Table 1**, **Figure 2E**, **Figure S7**).

### Changes in the profile of non-daily smokers

**Table 2** summarises changes in the sociodemographic, vaping, drinking, and smoking profiles of non-daily smokers across the study period.

**Table 2.** Sociodemographic, drinking, vaping, and smoking profile of non-daily smokers in England.

|  | Year<sup>1</sup> |  |  |  |  |  |
| --- | --- | --- | --- | --- | --- | --- |
|  | 2006-09 | 2009-12 | 2012-15 | 2015-18 | 2018-21 | 2021-24 |
| *Unweighted N* | 1,438 | 1,410 | 1,340 | 1,397 | 1,499 | 1,650 |
| Age (years) |  |  |  |  |  |  |
| Mean [95% CI] | 38.0 [37.1–38.8] | 38.6 [37.7–39.4] | 38.4 [37.5–39.2] | 37.5 [36.6–38.3] | 36.6 [35.8–37.4] | 36.5 [35.7–37.2] |
| 18-24 | 23.5 [21.2–26.1] | 20.6 [18.3–23.1] | 21.9 [19.8–24.3] | 26.3 [24.0–28.8] | 26.3 [23.9–28.7] | 27.1 [24.8–29.5] |
| 25-34 | 25.0 [22.7–27.5] | 26.2 [23.8–28.8] | 25.9 [23.4–28.6] | 26.4 [23.9–29.0] | 28.4 [25.9–31.1] | 29.7 [27.3–32.2] |
| 35-44 | 21.6 [19.4–24.0] | 22.5 [20.2–25.1] | 19.6 [17.3–22.1] | 15.6 [13.6–17.8] | 18.9 [16.8–21.2] | 16.3 [14.5–18.3] |
| 45-54 | 14.4 [12.6–16.4] | 13.6 [11.7–15.6] | 16.2 [14.1–18.6] | 15.5 [13.6–17.6] | 12.5 [10.9–14.4] | 12.1 [10.5–13.8] |
| 55-64 | 7.9 [6.6–9.5] | 10.0 [8.4–11.9] | 9.7 [8.2–11.5] | 9.1 [7.7–10.7] | 7.7 [6.5–9.2] | 8.7 [7.5–10.2] |
| ≥65 | 7.5 [6.2–9.0] | 7.1 [5.9–8.5] | 6.6 [5.4–8.1] | 7.1 [6.0–8.5] | 6.2 [5.1–7.5] | 6.1 [5.0–7.4] |
| Women | 48.3 [45.5–51.1] | 45.8 [43.0–48.6] | 46.5 [43.6–49.4] | 47.8 [45.1–50.6] | 42.9 [40.2–45.6] | 44.4 [41.7–47.0] |
| Social grade C2DE (less advantaged) | 45.4 [42.6–48.1] | 47.5 [44.7–50.3] | 44.6 [41.7–47.5] | 40.0 [37.3–42.9] | 47.9 [45.1–50.7] | 47.5 [44.9–50.2] |
| Level of alcohol consumption (AUDIT-C score)<sup>2,3</sup> |  |  |  |  |  |  |
| Mean (SD) | - | - | 4.5 [4.2–4.8] | 4.5 [4.3–4.7] | 4.5 [4.3–4.7] | 4.7 [4.6–4.9] |
| 0 (non-drinker) | - | - | 17.7 [15.0–20.8] | 19.0 [16.9–21.3] | 18.5 [16.4–20.8] | 17.3 [15.3–19.5] |
| 1-4 (low-risk) | - | - | 32.4 [28.5–36.4] | 30.3 [27.8–33.0] | 31.0 [28.5–33.7] | 28.9 [26.5–31.4] |
| 5-12 (increasing/higher-risk) | - | - | 49.9 [45.7–54.1] | 50.6 [47.8–53.4] | 50.5 [47.7–53.3] | 53.8 [51.1–56.5] |
| Current vaper<sup>3</sup> | - | 2.1 [1.3–3.4] | 19.8 [17.5–22.4] | 18.0 [16.0–20.2] | 23.2 [20.9–25.6] | 37.0 [34.5–39.5] |
| Cigarettes smoked per week, mean [95% CI] | 34.3 [32.2–36.4] | 36.7 [34.1–39.2] | 28.8 [26.6–30.9] | 24.7 [22.7–26.7] | 22.8 [21.1–24.6] | 21.1 [19.6–22.5] |
| Mainly/exclusively smokes hand-rolled cigarettes<sup>3</sup> | 22.7 [19.7–26.0] | 26.2 [23.7–28.8] | 33.6 [30.8–36.5] | 41.3 [38.5–44.2] | 41.3 [38.5–44.2] | 43.8 [41.0–46.7] |

Table 2. *continued*
|  | Year<sup>1</sup> |  |  |  |  |  |
| --- | --- | --- | --- | --- | --- | --- |
|  | 2006-09 | 2009-12 | 2012-15 | 2015-18 | 2018-21 | 2021-24 |
| Strength of urges to smoke<sup>4</sup> |  |  |  |  |  |  |
| Mean [95% CI] | 1.3 [1.3–1.4] | 1.3 [1.2–1.3] | 1.2 [1.1–1.2] | 1.1 [1.0–1.1] | 1.0 [0.9–1.1] | 1.1 [1.0–1.2] |
| 0. Not at all | 29.2 [26.7–31.9] | 30.9 [28.2–33.7] | 35.3 [32.5–38.3] | 41.4 [38.7–44.2] | 40.6 [37.9–43.3] | 38.0 [35.5–40.7] |
| 1. Slight | 24.0 [21.7–26.5] | 25.9 [23.3–28.6] | 26.5 [23.8–29.3] | 23.6 [21.3–26.0] | 28.4 [25.9–31.0] | 27.3 [25.0–29.7] |
| 2. Moderate | 36.2 [33.6–38.9] | 32.4 [29.6–35.2] | 28.0 [25.3–30.8] | 26.1 [23.8–28.7] | 24.1 [21.8–26.6] | 25.1 [22.9–27.5] |
| 3. Strong | 8.0 [6.6–9.6] | 8.2 [6.7–10.1] | 7.5 [6.1–9.2] | 6.1 [4.8–7.6] | 4.5 [3.5–5.8] | 6.8 [5.6–8.3] |
| 4. Very strong | 1.8 [1.2–2.7] | 1.7 [1.1–2.7] | 2.4 [1.6–3.6] | 2.0 [1.4–3.0] | 1.6 [1.0–2.5] | 2.2 [1.6–3.1] |
| 5. Extremely strong | 0.8 [0.4–1.4] | 0.9 [0.5–1.6] | 0.4 [0.1–1.0] | 0.8 [0.4–1.4] | 0.8 [0.4–1.6] | 0.5 [0.2–1.1] |
| Motivation to stop smoking<sup>3</sup> |  |  |  |  |  |  |
| Mean [95% CI] | 4.2 [4.0–4.4] | 4.1 [3.9–4.2] | 3.6 [3.5–3.7] | 3.3 [3.2–3.4] | 3.5 [3.4–3.6] | 3.5 [3.4–3.6] |
| 1. I don't want to stop smoking | 15.0 [11.6–19.1] | 18.3 [16.2–20.6] | 23.6 [21.2–26.1] | 26.6 [24.2–29.2] | 22.0 [19.8–24.4] | 22.3 [20.2–24.7] |
| 2. I think I should stop smoking but don't really want to | 8.8 [6.4–12.0] | 10.6 [9.0–12.5] | 15.0 [13.0–17.3] | 15.8 [13.8–17.9] | 19.3 [17.2–21.6] | 17.3 [15.3–19.4] |
| 3. I want to stop smoking but haven't thought about when | 11.5 [8.7–15.2] | 9.3 [7.7–11.2] | 8.0 [6.6–9.8] | 11.5 [9.8–13.3] | 11.3 [9.6–13.2] | 13.7 [12.0–15.7] |
| 4. I really want to stop smoking but I don't know when I will | 15.7 [12.5–19.6] | 16.4 [14.4–18.6] | 22.2 [19.9–24.8] | 18.0 [16.0–20.3] | 14.9 [13.0–17.0] | 13.0 [11.3–14.9] |
| 5. I want to stop smoking and hope to soon | 18.3 [14.7–22.4] | 16.5 [14.6–18.7] | 9.3 [7.8–11.0] | 10.6 [9.0–12.4] | 10.6 [9.0–12.5] | 12.7 [11.0–14.6] |
| 6. I really want to stop smoking and intend to in the next 3 months | 14.5 [11.3–18.3] | 11.8 [10.1–13.9] | 8.1 [6.7–9.8] | 7.3 [6.0–9.0] | 10.7 [9.1–12.6] | 10.4 [8.9–12.1] |
| 7. I really want to stop smoking and intend to in the next month | 16.3 [13.0–20.2] | 17.0 [14.9–19.3] | 13.8 [11.9–15.9] | 10.2 [8.7–12.0] | 11.2 [9.5–13.1] | 10.6 [9.1–12.4] |
| Number of past-year quit attempts |  |  |  |  |  |  |
| 0 | 57.3 [54.5–60.0] | 65.0 [62.2–67.7] | 62.2 [59.3–65.0] | 66.6 [63.9–69.2] | 63.4 [60.6–66.1] | 64.3 [61.7–66.8] |
| 1 | 25.2 [22.8–27.7] | 19.7 [17.5–22.0] | 21.5 [19.2–24.0] | 17.5 [15.5–19.8] | 19.6 [17.4–22.0] | 18.1 [16.1–20.3] |
| ≥2 | 17.5 [15.5–19.7] | 15.3 [13.4–17.5] | 16.2 [14.2–18.5] | 15.8 [13.8–18.0] | 17.0 [14.9–19.3] | 17.6 [15.6–19.7] |

Data are shown as column percentages with 95% CI, unless otherwise specified.
<sup>1</sup> Years are coded from November to October (i.e., November 2006 to October 2009, November 2009 to October 2012, etc.). Note 2021-24 includes data from November 2021 to April 2024 only.
<sup>2</sup> AUDIT-C, range = 0–12.
<sup>3</sup> Data not collected in every wave: the type of cigarettes smoked was assessed from January 2008, motivation to stop smoking from November 2008, current vaping from April 2011, and alcohol consumption from March 2014.
<sup>5</sup> Strength of urges to smoke, range = 0–5. See **Table S3** for data stratified by vaping status.

There was an uncertain decline in the mean age of non-daily smokers (from 38.0 years in 2006-09 to 36.5 in 2021-24) and a substantial increase in the proportion who vaped (from 2.1% in 2009-12 to 37.0% in 2021-24). There were no notable changes in terms of gender, occupational social grade, or level of alcohol consumption.

The proportion of non-daily smokers who mainly/exclusively smoked hand-rolled cigarettes increased considerably (from 22.7% in 2006-09 to 43.8% in 2021-24). The mean number of cigarettes smoked fell by more than a third (from 34.3 to 21.1 cigarettes per week). Levels of cigarette dependence also appeared to decline. In 2006-09, non-daily smokers most commonly reported experiencing moderate urges to smoke over the past 24 hours (36.2%). By 2021-24, this proportion had decreased to 25.1%, and the most common response was that they experienced no urges to smoke at all (38.0%). The proportions reporting strong, very strong, or extremely strong urges to smoke were relatively similar over time (10.6% in 2006-09, 9.5% in 2021-24). Levels of cigarette dependence were consistently higher among non-daily smokers who vaped than those who did not but declined over time in both groups (**Table S3**).

Motivation and attempts to quit smoking also declined. There were decreases in the proportions who said they really wanted to stop smoking and intended to do so in the next month (from 16.3% in 2006-09 to 10.6% in 2021-24) or the next three months (from 14.5% to 10.4%) and increases in the proportions who said they did not want to stop smoking (from 15.0% to 22.3%) or that they thought they should stop but did not really want to (from 8.8% to 17.3%). The proportion who had made at least one serious attempt to quit in the past year fell from 42.7% to 35.7%.

## Discussion

Between 2006 and 2013, the proportion of cigarette smokers in England who smoked non-daily was relatively stable, at around one in ten. However, this number increased considerably over the following decade: by April 2024, more than one in four adult cigarette smokers in England said they did not smoke every day and this increasing trend showed no signs of stopping. The increase in non-daily vaping was particularly pronounced among younger adults and those who vaped. Over time, non-daily smokers were, on average, slightly younger, more likely to vape, and more likely to smoke hand-rolled cigarettes. They also reported smoking fewer cigarettes each week and weaker urges to smoke, on average, indicating lower levels of cigarette dependence. However, they also appeared to be less motivated to stop smoking, and less likely to attempt to quit, compared with earlier years.

Among adults, the prevalence of non-daily smoking was consistently higher across the study period among those who were younger, those who vaped, and those who drank more heavily; groups that tend to have higher rates of smoking in general.^41,42^ Among cigarette smokers, prevalence was higher among those who were younger, from more advantaged social grades, and those who drank alcohol (i.e., AUDIT-C > 0), consistent with previous studies.^16,22,25^ In more recent years, non-daily smoking was also more common among those who vaped: this may partly reflect age differences between vapers and non-vapers (vaping prevalence has increased much more rapidly among younger than older adults in recent years^43–45)^.

Key findings were that non-daily smoking has increased over the past decade (2014–2024) after having been stable for many years, and that this increase has been particularly pronounced at younger ages. A possible explanation is that vaping may have had direct or indirect effects on non-daily smoking. The timing of the rise in non-daily smoking we observed (i.e., since 2014) coincided with the period since vaping has become popular.^43,46^ Nicotine dependence may have been displaced from cigarettes to e-cigarettes, making it easier to reduce cigarette consumption and to have longer periods between cigarettes without experiencing symptoms of withdrawal. Young adults who have taken up smoking since vaping has become popular may also have different smoking norms, including more non-daily use.

Another explanation may be financial pressures: it has become increasingly expensive to smoke^47^ as a result of tax increases on cigarettes and, in more recent years, financial impacts of the Covid-19 pandemic and cost-of-living crisis. Likely as a result of this, a growing proportion of people who smoke cigarettes (either daily or non-daily) have opted to use cheaper hand-rolled tobacco over manufactured cigarettes in recent years.^10^ On average, younger adults tend to have lower disposable incomes^48^ and those who smoke are less dependent on nicotine^49,50^ than at older ages. Younger smokers may therefore be more likely to adapt to the rising cost of smoking and limited budgets by not smoking every day. Further research (e.g., qualitative) would be useful to better understand changing patterns of non-daily smoking and subgroup differences.

Our findings have implications for public health policy. The UK Government recently committed to investing in mass media campaigns and stop smoking services to deter uptake of smoking and help existing smokers to quit. Non-daily smokers may be an important target as they represent a substantial and growing proportion of smokers who may underestimate the harms of their smoking^14,15^ and are decreasingly motivated to quit. Messaging could emphasise that even non-daily smoking is harmful to health^5–7,9^ and can be difficult to quit^21,22^ and encourage all smokers to use effective forms of support to boost their chances of success.

Key strengths of this study include the large, representative sample and monthly data collection over a period of 17.5 years. There were also several limitations. There was a change in mode of data collection during the study period, but when we collected data via both modalities (face-to-face and telephone) in the same month, estimates of smoking status were very similar.^36^ Vaping status, alcohol consumption, and the main type of cigarettes smoked were not assessed across the entire period. However, we note that vaping prevalence was very low before we started collecting data on this,^51^ so it would not have been possible to model trends in non-daily smoking by vaping status over a much longer period. Additionally, we did not have information on whether participants who had successfully stopped smoking the past year had previously been daily or non-daily smokers, which meant our analyses focused on current non-daily smoking. As a result, our quit attempts outcome only captured failed quit attempts. While this may affect absolute estimates of the proportion who tried to quit, it should not affect changes over time (given the same limitation applies across the study period).

## Conclusions

An increasing proportion of adults in England who smoke cigarettes do not smoke every day, particularly younger adults. Although non-daily smokers report smoking fewer cigarettes and weaker urges to smoke than they used to, which may make it easier for them to stop smoking, they appear to be decreasingly motivated to quit.

## Data Availability

All data produced in the present study are available upon reasonable request to the authors.

## List of abbreviations

AIC: Akaike Information Criterion
AUDIT-C: Alcohol Use Disorders Identification Test – Consumption scale
CI: confidence interval
PR: prevalence ratio

## Declarations

## Acknowledgments

Not applicable.

## Consent for publication

Not applicable.

## Authors’ contributions

Conceptualisation: SJ, SC, LS, JB. Data curation: JB. Formal analysis: SJ. Funding acquisition: LS, JB. Investigation: SJ, SC, LS, SCJB Methodology: SJ, SC, LS, JB. Supervision: JB. Visualisation: SJ. Writing – original draft: SJ. Writing – review & editing: SJ, SC, LS, JB. All authors read and approved the final manuscript.

## Ethics approval and consent to participate

Ethical approval for the STS was granted originally by the UCL Ethics Committee (ID 0498/001). The data are not collected by UCL and are anonymized when received by UCL. All participants provided verbal consent which was recorded on computers by trained interviewers.

## Authors’ Twitter handles

@DrSarahEJackson, @Sharon_Acox, @LionShahab, @jamiebrown10,

## References

1 Banks E, Joshy G, Weber MF, Liu B, Grenfell R, Egger S et al. Tobacco smoking and all-cause mortality in a large Australian cohort study: findings from a mature epidemic with current low smoking prevalence. BMC Med 2015; 13: 38.

2 Pirie K, Peto R, Reeves GK, Green J, Beral V. The 21st century hazards of smoking and benefits of stopping: a prospective study of one million women in the UK. The Lancet 2013; 381: 133–141.

3 Doll R, Peto R, Boreham J, Sutherland I. Mortality in relation to smoking: 50 years’ observations on male British doctors. BMJ 2004; 328: 1519.

4 Office of the Surgeon General (US), Office on Smoking and Health (US). The Health Consequences of Smoking: A Report of the Surgeon General. Centers for Disease Control and Prevention (US): Atlanta (GA), 2004 http://www.ncbi.nlm.nih.gov/books/NBK44695/ (accessed 7 Aug2018).

5 Inoue-Choi M, McNeel TS, Hartge P, Caporaso NE, Graubard BI, Freedman ND. Non-Daily Cigarette Smokers: Mortality Risks in the U.S. Am J Prev Med 2019; 56: 27–37.

6 Løchen M-L, Gram IT, Mannsverk J, Mathiesen EB, Njølstad I, Schirmer H et al. Association of occasional smoking with total mortality in the population-based Tromsø study, 2001–2015. BMJ Open 2017; 7: e019107.

7 Luoto R, Uutela A, Puska P. Occasional smoking increases total and cardiovascular mortality among men. Nicotine Tob Res Off J Soc Res Nicotine Tob 2000; 2: 133–139.

8 Bjerregaard BK, Raaschou-Nielsen O, Sørensen M, Frederiksen K, Tjønneland A, Rohrmann S et al. The effect of occasional smoking on smoking-related cancers. Cancer Causes Control 2006; 17: 1305–1309.

9 Schane RE, Ling PM, Glantz SA. Health Effects of Light and Intermittent Smoking. Circulation 2010; 121: 1518–1522.

10 Jackson SE, Tattan-Birch H, Buss V, Shahab L, Brown J. Trends in Daily Cigarette Consumption Among Smokers: A Population Study in England, 2008–2023. Nicotine Tob Res 2024; : ntae071.

11 Shiffman S, Paty JA, Kassel JD, Gnys M, Zettler-Segal M. Smoking behavior and smoking history of tobacco chippers. Exp Clin Psychopharmacol 1994; 2: 126–142.

12 Garnett C, Tombor I, Beard E, Jackson SE, West R, Brown J. Changes in smoker characteristics in England between 2008 and 2017. Addiction 2020; 115: 748–756.

13 Jamal A, Phillips E, Gentzke AS, Homa DM, Babb SD, King BA et al. Current Cigarette Smoking Among Adults - United States, 2016. MMWR Morb Mortal Wkly Rep 2018; 67: 53–59.

14 Ames S, Stevens S, Schroeder D, Werch C, Carlson J, Kiros G-E et al. Nondaily tobacco use among Black and White college undergraduates: A comparison of nondaily versus daily tobacco users. Addict Res Theory 2009; 17: 191–204.

15 Cooper TV, Taylor T, Murray A, DeBon MW, Vander Weg MW, Klesges RC et al. Differences between intermittent and light daily smokers in a population of U.S. military recruits. Nicotine Tob Res 2010; 12: 465–473.

16 Pinsker EA, Berg CJ, Nehl EJ, Prokhorov AV, Buchanan TS, Ahluwalia JS. Intent to quit among daily and non-daily college student smokers. Health Educ Res 2013; 28: 313–325.

17 Robertson L, Iosua E, McGee R, Hancox RJ. Nondaily, Low-Rate Daily, and High-Rate Daily Smoking in Young Adults: A 17-Year Follow-Up. Nicotine Tob Res 2016; 18: 943–949.

18 Berg CJ, Schauer GL, Buchanan TS, Sterling K, DeSisto C, Pinsker EA et al. Perceptions of addiction, attempts to quit, and successful quitting in nondaily and daily smokers. Psychol Addict Behav 2013; 27: 1059–1067.

19 DiFranza JR, Savageau JA, Fletcher K, Ockene JK, Rigotti NA, McNeill AD et al. Measuring the loss of autonomy over nicotine use in adolescents: the DANDY (Development and Assessment of Nicotine Dependence in Youths) study. Arch Pediatr Adolesc Med 2002; 156: 397–403.

20 Ursprung WWSA, DiFranza JR. The loss of autonomy over smoking in relation to lifetime cigarette consumption. Addict Behav 2010; 35: 14–18.

21 Tindle HA, Shiffman S. Smoking cessation behavior among intermittent smokers versus daily smokers. Am J Public Health 2011; 101: e1–3.

22 Kotz D, Fidler J, West R. Very low rate and light smokers: smoking patterns and cessation-related behaviour in England, 2006–11. Addiction 2012; 107: 995–1002.

23 Omole T, McNeel T, Choi K. Heterogeneity in past-year smoking, current tobacco use, and smoking cessation behaviors among light and/or non-daily smokers. Tob Induc Dis 2020; 18: 74.

24 Tong EK, Ong MK, Vittinghoff E, Pérez-Stable EJ. Nondaily smokers should be asked and advised to quit. Am J Prev Med 2006; 30. doi:10.1016/j.amepre.2005.08.048.

25 Wortley PM, Husten CG, Trosclair A, Chrismon J, Pederson LL. Nondaily smokers: a descriptive analysis. Nicotine Tob Res Off J Soc Res Nicotine Tob 2003; 5: 755–759.

26 Harrison ELR, McKee SA. Young adult non-daily smokers: Patterns of alcohol and cigarette use. Addict Behav 2008; 33: 668–674.

27 Harrison ELR, Desai RA, McKee SA. Nondaily smoking and alcohol use, hazardous drinking, and alcohol diagnoses among young adults: findings from the NESARC. Alcohol Clin Exp Res 2008; 32: 2081–2087.

28 White HR, Bray BC, Fleming CB, Catalano RF. Transitions into and out of light and intermittent smoking during emerging adulthood. Nicotine Tob Res Off J Soc Res Nicotine Tob 2009; 11: 211–219.

29 McKee SA, Hinson R, Rounsaville D, Petrelli P. Survey of subjective effects of smoking while drinking among college students. Nicotine Tob Res Off J Soc Res Nicotine Tob 2004; 6: 111–117.

30 Coleman SRM, Piper ME, Byron MJ, Bold KW. Dual Use of Combustible Cigarettes and E-cigarettes: a Narrative Review of Current Evidence. Curr Addict Rep 2022; 9: 353–362.

31 McNeill A, Simonavicius E, Brose LS, Taylor E, East K, Zuikova E et al. Nicotine vaping in England: an evidence update including health risks and perceptions, September 2022. A report commissioned by the Office for Health Improvement and Disparities. Office for Health Improvement and Disparities: London, 2022https://www.gov.uk/government/publications/nicotine-vaping-in-england-2022-evidence-update (accessed 3 Oct2022).

32 Lindson N, Butler AR, McRobbie H, Bullen C, Hajek P, Begh R et al. Electronic cigarettes for smoking cessation. Cochrane Database Syst Rev 2024. doi:10.1002/14651858.CD010216.pub8.

33 Fidler JA, Shahab L, West O, Jarvis MJ, McEwen A, Stapleton JA et al. ‘The smoking toolkit study’: a national study of smoking and smoking cessation in England. BMC Public Health 2011; 11: 479.

34 Kock L, Shahab L, Moore G, Beard E, Bauld L, Reid G, et al. Protocol for expansion of an existing national monthly survey of smoking behaviour and alcohol use in England to Scotland and Wales: The Smoking and Alcohol Toolkit Study. Wellcome Open Res 2021; 6: 67.

35 Jackson SE, Beard E, Kujawski B, Sunyer E, Michie S, Shahab L et al. Comparison of Trends in Self-reported Cigarette Consumption and Sales in England, 2011 to 2018. JAMA Netw Open 2019; 2: e1910161.

36 Kock L, Tattan-Birch H, Jackson S, Shahab L, Brown J. Socio-demographic, smoking and drinking characteristics in GB: A comparison of independent telephone and face-to-face Smoking and Alcohol Toolkit surveys conducted in March 2022. Qeios 2022. doi:10.32388/CLXK4D.

37 Jackson SE, Garnett C, Shahab L, Oldham M, Brown J. Association of the Covid-19 lockdown with smoking, drinking, and attempts to quit in England: an analysis of 2019-2020 data. Addiction 2021; 116: 1233–1244.

38 Babor TF, Higgins-Biddle JC, Saunders JB, Monteiro MG, Organization WH. AUDIT: the alcohol use disorders identification test: guidelines for use in primary health care. 2001.

39 Fidler JA, Shahab L, West R. Strength of urges to smoke as a measure of severity of cigarette dependence: comparison with the Fagerström Test for Nicotine Dependence and its components. Addict Abingdon Engl 2011; 106: 631–638.

40 Kotz D, Brown J, West R. Predictive validity of the Motivation To Stop Scale (MTSS): A single-item measure of motivation to stop smoking. Drug Alcohol Depend 2013; 128: 15–19.

41 Office for National Statistics. Adult smoking habits in the UK: 2022. 2023 https://www.ons.gov.uk/peoplepopulationandcommunity/healthandsocialcare/healthandlifeexpectancies/bulletins/adultsmokinghabitsingreatbritain/2022 (accessed 7 Sep2023).

42 Garnett C, Oldham M, Shahab L, Tattan-Birch H, Cox S. Characterising smoking and smoking cessation attempts by risk of alcohol dependence: A representative, cross-sectional study of adults in England between 2014-2021. Lancet Reg Health – Eur 2022; 18. doi:10.1016/j.lanepe.2022.100418.

43 Action on Smoking and Health. Use of e-cigarettes among adults in Great Britain. 2023 https://ash.org.uk/resources/view/use-of-e-cigarettes-among-adults-in-great-britain-2021 (accessed 30 Oct2023).

44 Jackson SE, Tattan-Birch H, Shahab L, Oldham M, Kale D, Brose L et al. Who would be affected by a ban on disposable vapes? A population study in Great Britain. Public Health 2024; 227: 291–298.

45 Tattan-Birch H, Brown J, Shahab L, Beard E, Jackson SE. Trends in vaping and smoking following the rise of disposable e-cigarettes: a repeat cross-sectional study in England between 2016 and 2023. Lancet Reg Health - Eur 2024; : 100924.

46 Jackson SE, Brown J, Beard E. Associations of Prevalence of E-cigarette Use With Quit Attempts, Quit Success, Use of Smoking Cessation Medication, and the Overall Quit Rate Among Smokers in England: A Time-Series Analysis of Population Trends 2007–2022. Nicotine Tob Res 2024; : ntae007.

47 Jackson SE, Tattan-Birch H, Shahab L, Brown J. How has Expenditure on Nicotine Products Changed in a Fast-Evolving Marketplace? A Representative Population Survey in England, 2018–2022. Nicotine Tob Res 2023; : ntad074.

48 Office for National Statistics. Household disposable income by age group. 2021.https://www.ons.gov.uk/aboutus/transparencyandgovernance/freedomofinformationfoi/householddisposableincomebyagegroup (accessed 2 Mar2023).

49 Schnoll RA, Goren A, Annunziata K, Suaya JA. The prevalence, predictors and associated health outcomes of high nicotine dependence using three measures among US smokers. Addiction 2013; 108: 1989–2000.

50 Goodwin RD, Pagura J, Spiwak R, Lemeshow AR, Sareen J. Predictors of persistent nicotine dependence among adults in the United States. Drug Alcohol Depend 2011; 118: 127–133.

51 Buss V, West R, Kock L, Kale D, Brown J. Monthly trends on smoking in England from the Smoking Toolkit Study. 2024.https://smokinginengland.info/graphs/monthly-tracking-kpi (accessed 8 Feb2023).

