## Supplementary file for "Trends in non-daily cigarette smoking in England, 2006–2024"

**Table S1. Model selection: AIC values for models with 3, 4, and 5 knots**

| | AIC | | |
|---|---|---|---|
| | 3 knots | 4 knots | 5 knots |
| Adults | | | |
| Overall | 84094.21 | 84096.22 | 84098.73 |
| By age | 81001.88 | 81005.57 | 81002.81 |
| By gender | 83563.32 | 83566.09 | 83568.33 |
| By social grade | 84081.01 | 84076.38 | 84053.82 |
| By vaping status | 79759.90 | 79760.93 | 79734.71 |
| By alcohol consumption | 47506.86 | 47505.12 | 47509.13 |
| Cigarette smokers | | | |
| Overall | 52417.53 | 52419.16 | 52421.58 |
| By age | 51445.00 | 51447.41 | 51450.59 |
| By gender | 52180.45 | 52184.20 | 52188.67 |
| By social grade | 51455.64 | 51434.93 | 51433.83 |
| By vaping status | 50833.76 | 50835.59 | 50829.92 |
| By alcohol consumption | 28024.34 | 28021.74 | 28019.87 |

AIC, Akaike Information Criterion.

Shaded cells indicate the best fitting model (the model with the lowest AIC or the simplest model within 2 AIC units).

**Table S2. Modelled estimates within age groups of the prevalence of non-daily smoking among adults in the first available monthly wave and in April 2024**

| | Adults | | Cigarette smokers | |
|---|---|---|---|---|
| | Prevalence, % [95%CI][1] | | Prevalence, % [95%CI][1] | |
| | Nov 2006 | Apr 2024 | Nov 2006 | Apr 2024 |
| Age group (years) | | | | |
| 18–24 | 4.6 [4.1–5.1] | 10.1 [9.3–11.1] | 14.7 [13.3–16.4] | 48.2 [45.3–51.1] |
| 25–34 | 3.8 [3.5–4.1] | 6.9 [6.5–7.3] | 12.5 [11.6–13.5] | 36.0 [34.2–37.8] |
| 35–44 | 3.0 [2.7–3.2] | 4.3 [4.0–4.7] | 10.5 [9.6–11.4] | 24.8 [23.2–26.5] |
| 45–54 | 2.2 [2.0–2.5] | 2.8 [2.5–3.0] | 9.2 [8.3–10.1] | 18.2 [16.7–19.7] |
| 55–64 | 1.6 [1.4–1.8] | 1.8 [1.7–2.0] | 8.5 [7.7–9.5] | 15.2 [13.9–16.5] |
| ≥65 | 0.8 [0.6–1.0] | 0.9 [0.8–1.2] | 8.3 [6.3–10.9] | 14.1 [11.3–17.5] |

[1] Data are weighted estimates of prevalence in the first and last months in the study period from logistic regression with survey month and age modelled non-linearly using restricted cubic splines. They are calculated as means of modelled estimates across years of age in each age group (e.g., prevalence among 18-24-year-olds was calculated as the mean of modelled estimates for those aged 18, 19, 20, 21, 22, 23, and 24 years).

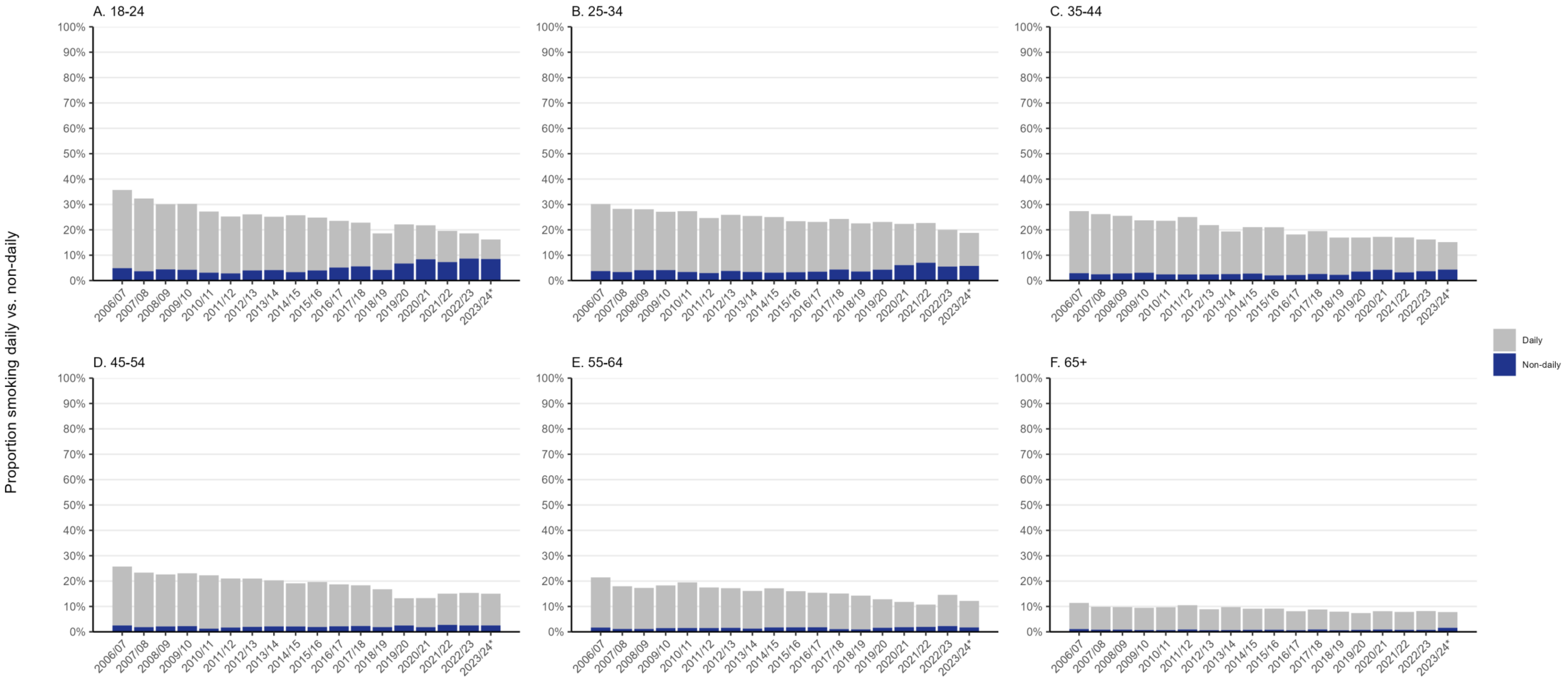


**Figure S1. Prevalence of daily and non-daily smoking, by age.**

Panels show weighted data aggregated by year among adults in England. Bars represent the proportions smoking daily and non-daily. Data are aggregated across 12-month periods (November to October). *Data for 2023/24 are based on November to April only.

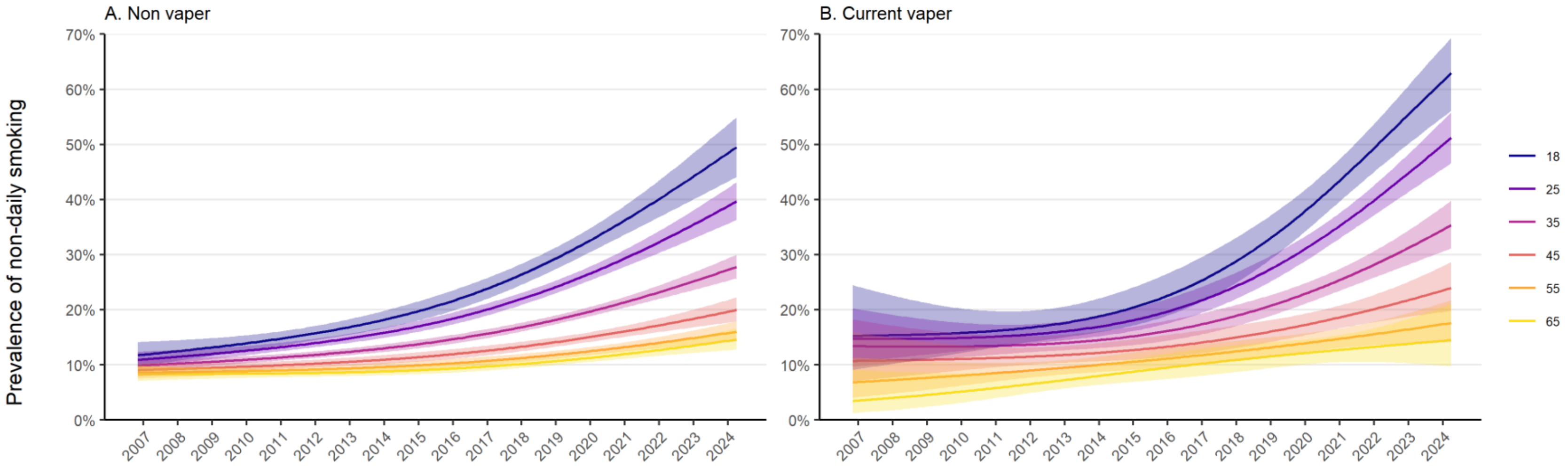


**Figure S2. Trends by age in non-daily smoking among adult cigarette smokers in England, stratified by vaping status.**

Lines represent the modelled weighted proportion by monthly survey wave (modelled non-linearly using restricted cubic splines; three knots) and age, stratified by vaping status. Shaded bands represent 95% confidence intervals. Note: data on vaping status were only available for cigarette smokers from April 2011.

Unweighted sample sizes: non-vapers *n*=34,566; current vapers *n*=7,693.

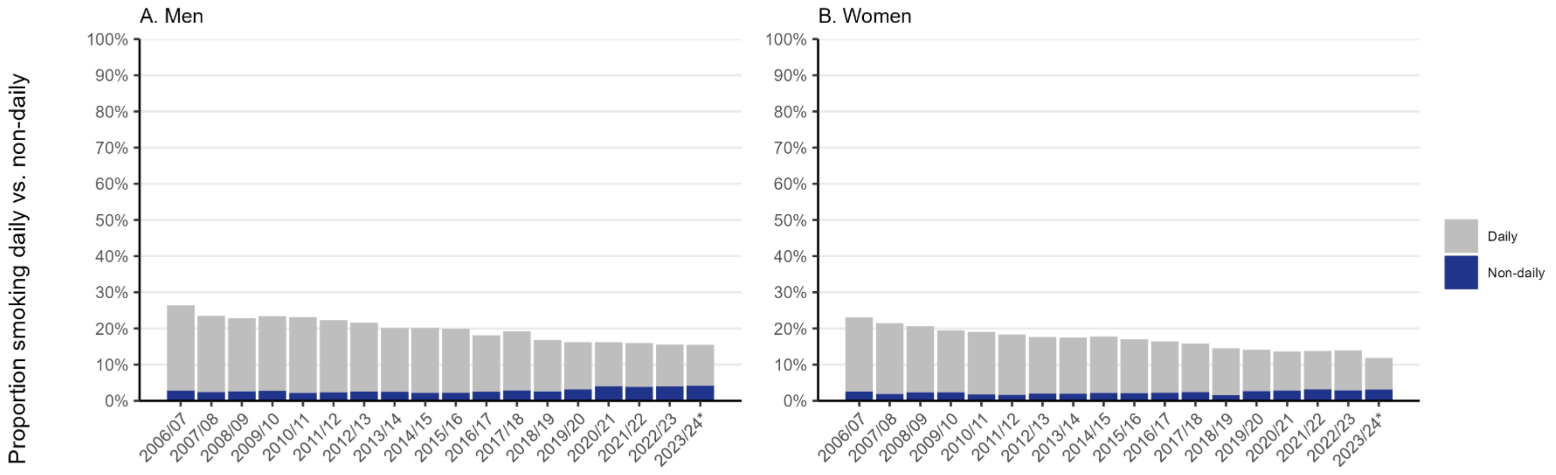


**Figure S3. Prevalence of daily and non-daily smoking, by gender.**

Panels show weighted data aggregated by year among adults in England. Bars represent the proportions smoking daily and non-daily. Data are aggregated across 12-month periods (November to October). *Data for 2023/24 are based on November to April only.

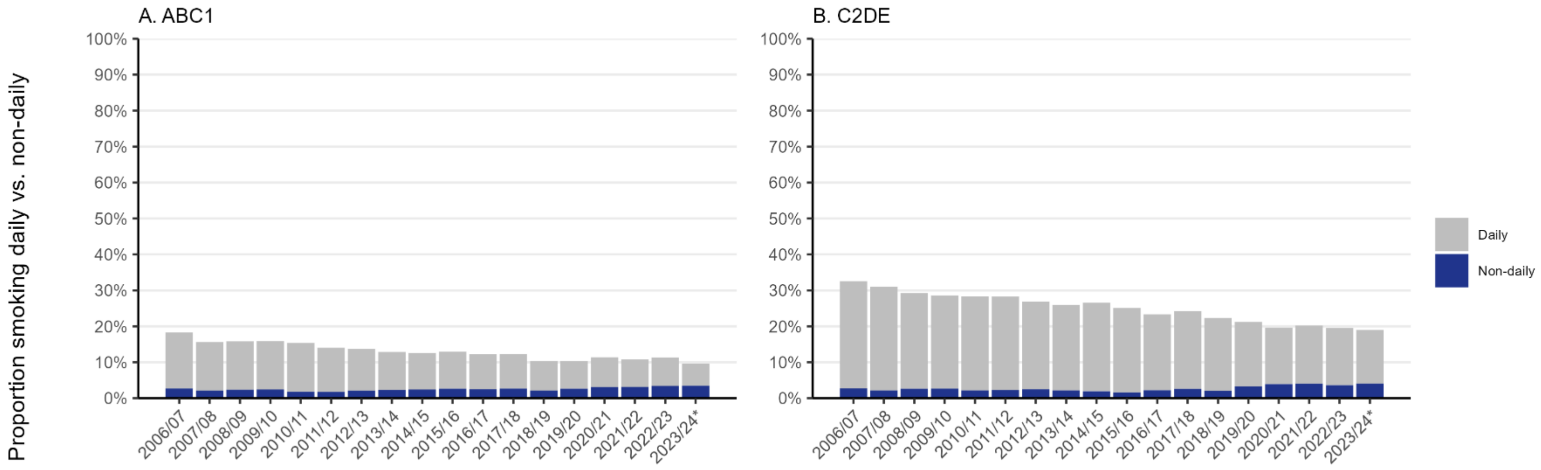


**Figure S4. Prevalence of daily and non-daily smoking, by occupational social grade.**

Panels show weighted data aggregated by year among adults in England. Bars represent the proportions smoking daily and non-daily. Data are aggregated across 12-month periods (November to October). *Data for 2023/24 are based on November to April only.

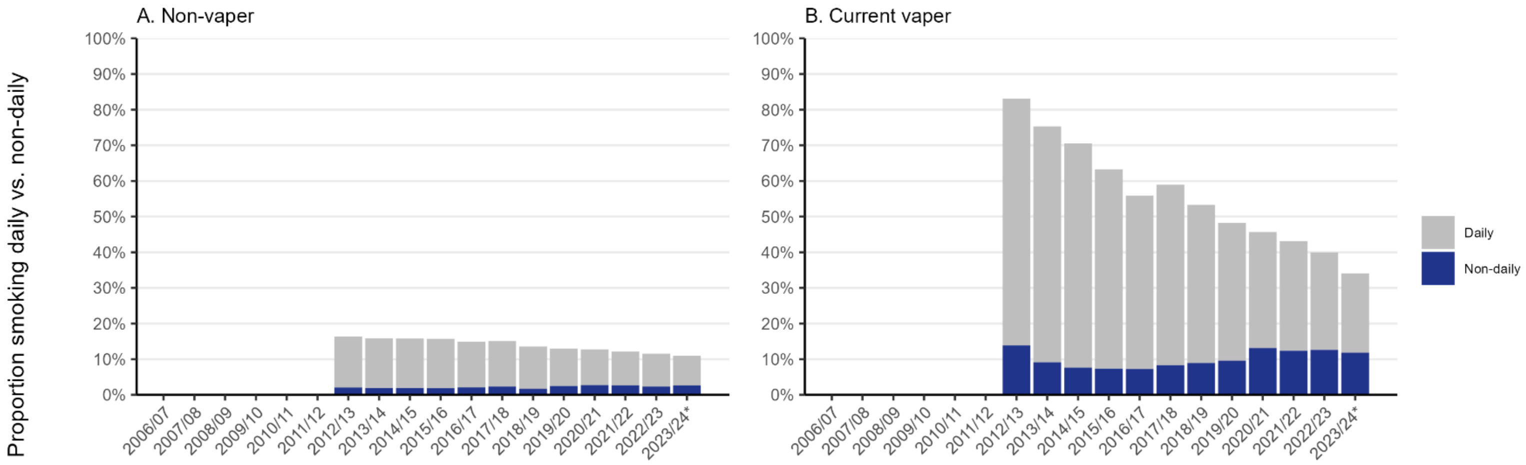


**Figure S5. Prevalence of daily and non-daily smoking, by vaping status.**

Panels show weighted data aggregated by year among adults in England. Bars represent the proportions smoking daily and non-daily. Data are aggregated across 12-month periods (November to October). Data on vaping status were collected from October 2013. *Data for 2023/24 are based on November to April only.

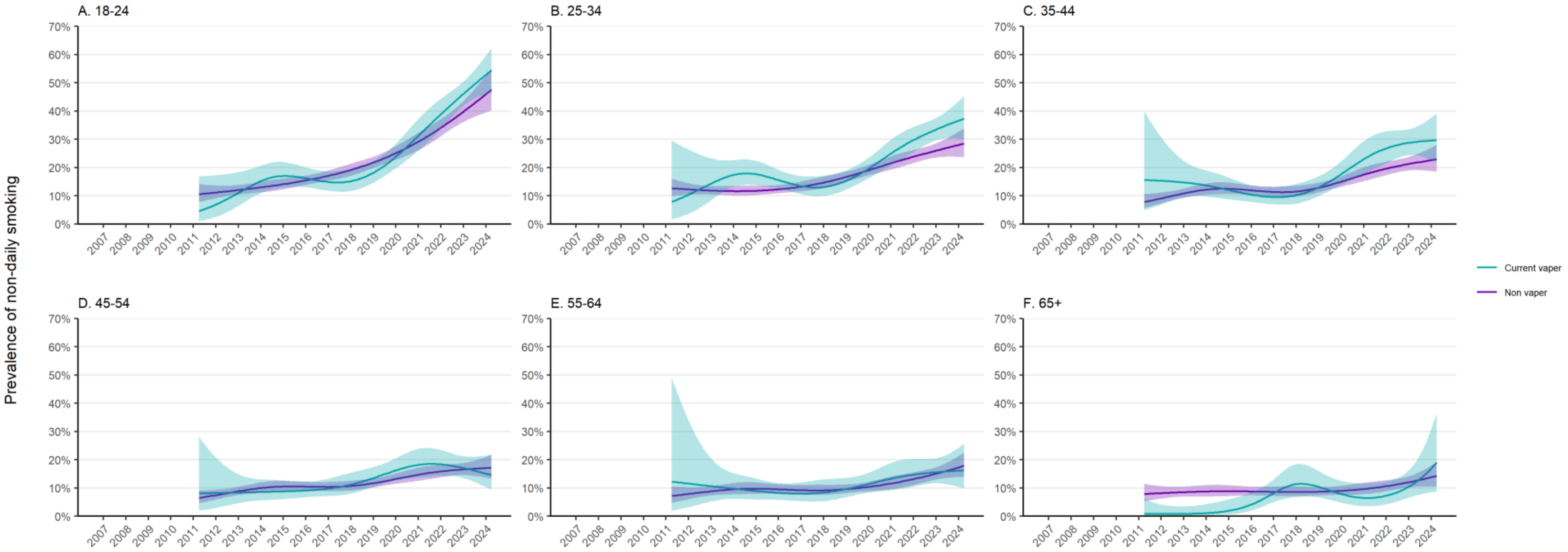


**Figure S6. Trends by vaping status in non-daily smoking among adult cigarette smokers in England, stratified by age.**

Lines represent the modelled weighted proportion by monthly survey wave (modelled non-linearly using restricted cubic splines; five knots) and vaping status, stratified by age group. Shaded bands represent 95% confidence intervals. Note: data on vaping status were only available for cigarette smokers from April 2011.

Unweighted sample sizes: 18-24 *n*=7,234; 25-34 *n*=8,882; 35-44 *n*=7,161; 45-54 *n*=7,153; 55-64 *n*=6,106; 65+ *n*=5,714.

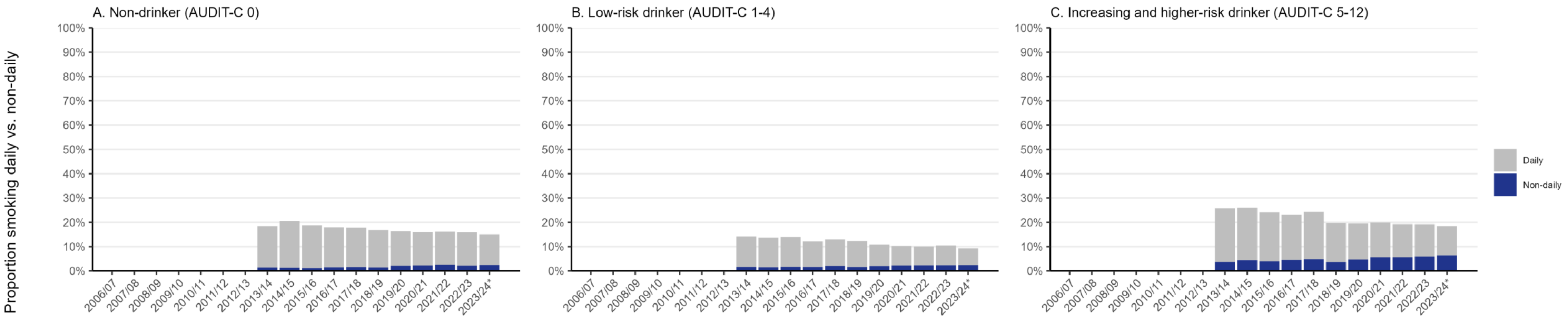


**Figure S7. Prevalence of daily and non-daily smoking, by level of alcohol consumption.**

Panels show weighted data aggregated by year among adults in England. Bars represent the proportions smoking daily and non-daily. Data are aggregated across 12-month periods (November to October). Data on alcohol consumption were collected from April 2014. *Data for 2023/24 are based on November to April only.

**Table S3.** Strength of urges to smoke among non-daily smokers in England, by vaping status

| | Year[1] | | | | | |
|---|---|---|---|---|---|---|
| | 2006-09 | 2009-12 | 2012-15 | 2015-18 | 2018-21 | 2021-24 |
| **Non vapers (exclusive smokers)** | | | | | | |
| Mean [95% CI] | - | 1.2 [1.2–1.3] | 1.1 [1.0–1.1] | 0.9 [0.9–1.0] | 0.9 [0.8–1.0] | 0.9 [0.8–1.0] |
| 0. Not at all | - | 31.6 [28.8–34.5] | 38.8 [35.5–42.2] | 46.9 [43.8–50.0] | 45.7 [42.6–48.9] | 46.1 [42.8–49.5] |
| 1. Slight | - | 26.2 [23.6–28.9] | 25.9 [23.0–29.1] | 23.5 [21.0–26.2] | 27.6 [24.8–30.5] | 25.7 [22.9–28.7] |
| 2. Moderate | - | 31.6 [28.8–34.4] | 26.3 [23.4–29.4] | 22.9 [20.4–25.6] | 20.3 [17.9–22.9] | 21.7 [19.0–24.6] |
| 3. Strong | - | 8.0 [6.5–9.8] | 6.9 [5.4–8.7] | 4.4 [3.2–6.0] | 4.2 [3.1–5.7] | 3.8 [2.8–5.3] |
| 4. Very strong | - | 1.7 [1.1–2.7] | 2.1 [1.3–3.3] | 1.7 [1.1–2.7] | 1.4 [0.9–2.4] | 2.2 [1.4–3.5] |
| 5. Extremely strong | - | 0.9 [0.5–1.7] | 0.1 [0.0–0.4] | 0.6 [0.3–1.3] | 0.7 [0.3–1.7] | 0.4 [0.2–1.2] |
| **Current vapers (dual users)** | | | | | | |
| Mean [95% CI] | - | 2.1 [1.8–2.4] | 1.5 [1.3–1.7] | 1.7 [1.5–1.8] | 1.4 [1.2–1.5] | 1.4 [1.3–1.5] |
| 0. Not at all | - | 21.4 [16.3–27.5] | 16.6 [12.4–22.0] | 23.5 [18.8–28.8] | 24.4 [20.9–28.3] | 11.4 [3.1–33.7] |
| 1. Slight | - | 28.7 [22.9–35.3] | 23.8 [18.6–29.9] | 31.1 [25.9–36.9] | 30.0 [26.1–34.2] | 68.4 [43.7–85.8] |
| 2. Moderate | - | 34.9 [28.6–41.8] | 40.8 [34.5–47.3] | 36.6 [31.1–42.6] | 30.9 [27.0–35.0] | 17.7 [5.7–43.2] |
| 3. Strong | - | 9.9 [6.6–14.6] | 13.9 [10.1–18.9] | 5.5 [3.5–8.5] | 11.8 [9.2–15.1] | 2.6 [0.3–19.3] |
| 4. Very strong | - | 3.7 [1.7–8.2] | 3.5 [1.8–7.0] | 2.2 [0.9–5.1] | 2.2 [1.3–3.7] | 1.4 [0.4–4.8] |
| 5. Extremely strong | - | 1.3 [0.5–3.6] | 1.2 [0.4–3.3] | 0.7 [0.2–2.3] | 1.2 [1.2–1.3] | 1.1 [1.0–1.1] |

Data are shown as column percentages with 95% CI, unless otherwise specified.

[1] Years are coded from November to October (i.e., November 2009 to October 2012, etc.). Note 2021-24 includes data from November 2021 to April 2024 only. No data are provided for 2006-2009 because vaping status was not assessed before April 2011.
